# A distinct symptom pattern emerges for COVID-19 Long-Haul: A nationwide study

**DOI:** 10.1101/2022.07.21.22277910

**Authors:** Charles A Downs, Melissa D. Pinto, Yong Huang, Survivor Corps, Sarah A. El-Azab, Nathan S. Ramrakhiani, Anthony Barisano, Lu Yu, Kaitlyn Taylor, Alvaro Esperanca, Heather L. Abrahim, Thomas Hughes, Maria Giraldo Herrera, Amir M. Rahmani, Nikil Dutt, Rana Chakraborty, Christian Mendiola, Natalie Lambert

**Affiliations:** School of Nursing & Health Studies, University of Miami, Coral Gables, Fl; University of California Irvine, Sue & Bill Gross School of Nursing, Irvine California; University of California Irvine, Donald Bren School of Information and Computer Science, Department of Computer Science; University of Michigan, Department of Health Management and Policy; Brown University, Providence, RI; University of Illinois at Urbana-Champaign, School of Labor and Employment Relations; Indiana University School of Medicine, Lambert Health Lab; Indiana University Purdue University Indianapolis, Electrical and Computer Engineering Department; Mayo Clinic, Division of Infectious Disease, Department of Pediatrics and Adolescent Medicine, Rochester, Minnesota; Indiana University Purdue University Indianapolis, School of Engineering and Technology; Indiana University School of Medicine, Department of Biostatistics and Health Data Sciences

**Keywords:** COVID-long-haul, Symptom Clusters, Long COVID, COVID-19 pandemic, post-acute sequalae of SARS-CoV-2 (PASC)

## Abstract

Long-haul COVID-19, also called Post-Acute Sequelae of SARS-CoV-2 (PASC), is a new illness caused by SARS-CoV-2 infection and characterized by the persistence of symptoms. The purpose of this cross-sectional study was to identify a distinct and significant temporal pattern of PASC symptoms (symptom type and onset) among a nationwide sample of PASC survivors (n= 5,652). The sample was randomly sorted into two independent samples for exploratory (EFA) and confirmatory factor analyses (CFA). Five factors emerged from the EFA: (1) cold & flu-like symptoms, (2) change in smell and/or taste, (3) dyspnea and chest pain, (4) cognitive & visual problems, and (5) cardiac symptoms. The CFA had excellent model fit (*x*^2^ = 513.721, df= 207, p<0.01, TLI= 0.952, CFI= 0.964, RMSEA= 0.024). These findings demonstrate a novel symptom pattern for PASC. These findings can enable nurses in the identification of at-risk patients and facilitate early, systematic symptom management strategies for PASC.

## Introduction

Long-haul COVID-19, post-acute sequelae of SARS-CoV-2 (PASC), is the persistence of symptoms that extend beyond the expected resolution of illness. PASC often causes significant disability and given the large-scale of the pandemic coupled with the lack of treatments, PASC is a global health crisis (1, 2). PASC has not yet been clinically characterized. The incidence PASC is estimated 11% to 80% (3-8) Initial COVID-19 symptom presentation varies; however, the development of PASC appears to be independent of COVID-19 symptom presentation, severity, or the presence of pre-morbid (pre-existing) health conditions (9). Persons at risk for PASC include those who were initially asymptomatic, as well as symptomatic persons independent of needing hospitalization. (4, 10, 11). PASC survivors report a variety of persistent and distressing symptoms that can last anywhere from weeks to more than a year (9). Nearly two years into the pandemic, PASC survivors continue to report symptoms and it is unclear if symptoms will eventually resolve or if a new chronic disease has emerged. We and others have numerated and characterized PASC symptoms at the onset of illness. (4, 9, 10, 12-14). However, it is still not known how PASC symptoms evolve over time, how they temporally group or cluster, and if findings represent a statistically significant pattern.

PASC symptom presentation, and more specifically the order in which symptoms are experienced, is useful for developing and implementing self-management strategies to reduce symptom burden and improve quality of life and daily functioning. As experts in symptom science, nurse scientists are well-poised to investigate PASC symptoms and leverage the existing self-management evidence base to develop or re-tool interventions to mitigate the devastating consequences of PASC symptoms (9). Further, understanding the PASC symptom pattern allows for the exploration of the underlying biological mechanisms underpinning symptoms and their evolution (9). Understanding symptoms and their context (i.e., biology, behaviors, etc..) is critical for the development of new interventions and pharmacological and non-pharmacological therapeutics. This study extends beyond symptom description and advances our understanding of PASC symptoms through added context, specifically symptom onset. Therefore, the aim of this study was to identify a distinct and statistically significant pattern of PASC symptoms through assessment of symptom type and symptom onset in a nationwide sample of PASC survivors (n=5,562). PASC is operationalized as experiencing symptoms more than 28 days after SARS-CoV-2 infection. In this paper, we used exploratory factor analysis (EFA) and confirmatory factor analysis (CFA). EFA was used to identify the underlying PASC symptom structure (symptom type and onset) and the temporal clustering of symptoms. CFA was used to validate findings of the EFA PASC symptom structure, and thereby yielded a distinct, statistically significant temporal pattern of symptoms among PASC survivors.

## Methods

### Sample and Setting

A convenience sample (n=5,562) of COVID-19 survivors were recruited (August 2020 through February 2021) through online COVID-19 survivor groups and online COVID support communities in response to a written study advertisement. Since study recruitment and data collection were completed online, the sampling frame was national. Inclusion criteria were English speaking, adults at least 18 years of age or older, and either a (self-report) positive PCR or history of healthcare provider confirmed clinical diagnosis of SARS-CoV-2 infection.

### Survey and Procedures

Institutional review board (IRB) approval was obtained from Indiana University prior to recruitment and data collection. Informed consent as approved by the IRB was implied prior to the completion of the survey. Since there is no gold-standard symptom survey measure for PASC, a symptom survey was developed using unstructured social media data in which patients reported their symptoms in free text. Further details regarding survey development are reported in (Authors, in review) (14). The survey asked participants to indicate (check) symptoms they had experienced since the onset of COVID-19 and at the time of completing the survey. Participants were then asked to write in symptom onset (the number of days after SARS-CoV-2 infection that a symptom began) of symptoms they checked. A free text option was included in which participants could report any symptom not included in the survey. Symptoms reported in the free text were evaluated for consistency across participants and collapsed into common categories as appropriate. All data collection were completed electronically using REDCap.

### Analytic Plan

Statistical analyses were performed using R software (Version psych 2.1.6 (EFA) and Lavaan 0.6-9 (CFA)). To ensure group equivalence descriptive statistics and measures of central tendency and *t-test* were used to describe the sample, test for differences at baseline, and onset of symptoms.

A sample of 5,562 participants were recruited, and 5,136 met criteria. The sample (n=5,136) was randomly sorted into two independent samples for EFA (n= 2,547) and CFA (n = 2589) analyses using a random number generator. The sample size exceeded the minimum criteria for subjects per item (10:1) ratio needed to conduct rigorous exploratory and confirmatory factor analyses. The following criteria were used to guide the exploratory and confirmatory factor analyses:

#### Exploratory Factor Analysis

##### Extraction and Rotation Method

A series of EFAs were performed using R to determine the factor structure of symptom onset for PASC. Latent factor structure was assessed through principal axis factoring (PAF) to extract the symptom onset for PASC factor structure. PAF accounts for the unique contribution of each item and can robustly analyze the data that violate the assumption of normality. Prior to the EFA, indicators of sampling adequacy were verified by Kaiser-Meyer-Olkin (KMO) and Bartlett’s Test of Sphericity (15).

##### Determination of Factors and Item Reduction

The scree plot and eigenvalue values for each factor guided the evaluation to determine the most parsimonious factor structure. Criteria reported by Costello and Osborne were used for interpretation of the scree plot (15). Kaiser criterion, which recommends an eigenvalue ≥1, was used for factor retention and the assessment of eigenvalues (16). The interpretation of the scree plot and eigenvalue of each factor determined the number of factors captured by the symptom onset of PASC.

##### Item Retention and Removal Criteria

*A priori* criteria for item retention were had primary factor loadings ≥ 0.40, secondary factor loadings <0.30, and did not have primary factor loadings on more than one factor (17). Items not meeting these criteria were removed individually. The EFA was repeated until all retained items met these criteria.

##### Labeling of Factors and Internal Reliability Consistency

The parsimonious factor structure was derived from the EFA and then labeled based content of the items retained. Evaluation of the internal reliability consistency of each factor was performed to assess the contribution of each item to the factor or total score of the symptom onset for PASC.

### Confirmatory Factor Analysis

#### Determination of Model Fit

The symptom onset for PASC model identified by the EFA was evaluated for validity using a first-order CFA. We used the following goodness-of-fit indices to determine model fit: *x*^*2*^, Tucker Lewis Index (LTI: >0.90 acceptable, >0.95 excellent), the Comparative Fit Index (CFA: >0.90 acceptable, >0.95 excellent), and the Root Mean Square Error of Approximation (RMSEA: <0.08 acceptable, <0.05 excellent) (18). Paths between error terms were added to enhance the goodness-to-fit of these data to the model based on interpretation of the above modification indices (19).

## Results

### Sample Characteristics

Sample demographics are provided in Table 1. No differences were observed between the EFA, and CFA independent samples based on sex, race/ethnicity, and age. The average time from SARS-CoV-2 infection to completing the questionnaire was 105.2 days (SD 58).

### Exploratory Factor Analysis to Identify Underlying Factor Structure

The first independent sample of 2,547 PASC survivors was used for EFA. In the EFA, the most parsimonious structure included five factors with a variable number of items per factor (range 3-10). See Table 2 for factor loadings. The mean onset of each symptom and its corresponding factor are presented in Figure 1. Based on the content of the items retained on each factor and the magnitude of these items’ factor loading, each factor was labeled as follows: Cold & flu-like symptoms (Factor 1), Change in Smell and/or taste (Factor 2), Dyspnea & chest pain (Factor 3), Cognitive-visual symptoms (Factor 4), and Cardiac symptoms (Factor 5). For each factor the item-to-total correlations (range 0.50-0.95) and inter-item correlations (range (0.23-0.92)) are provided in Table 3. See Figure 2 for the EFA model.

**Figure 1:**
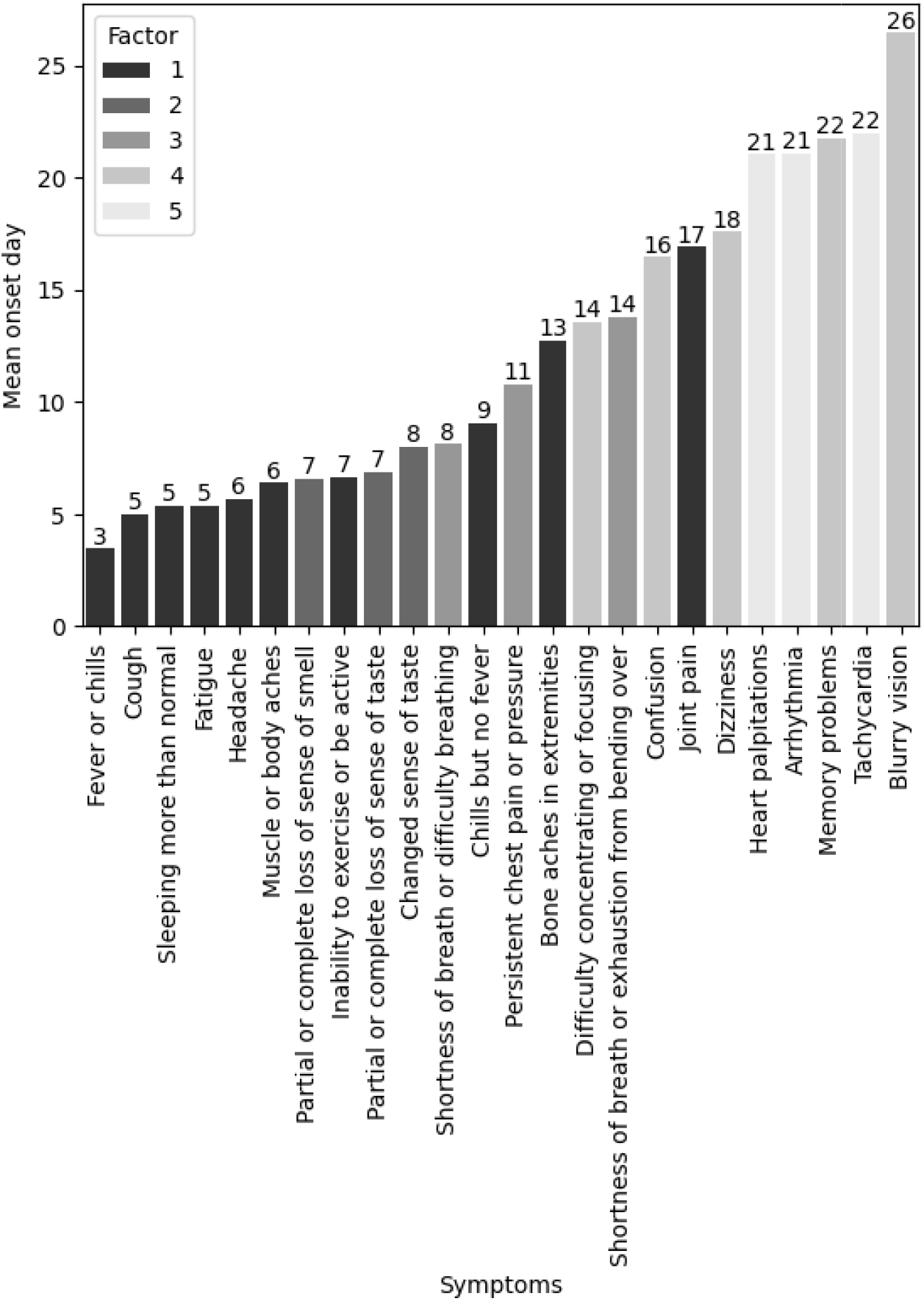
Mean Onset of Symptoms with Factor Loading. Bar graph with showing mean symptom onset (in days) and associated factor loading for each symptom.

**Figure 2:**
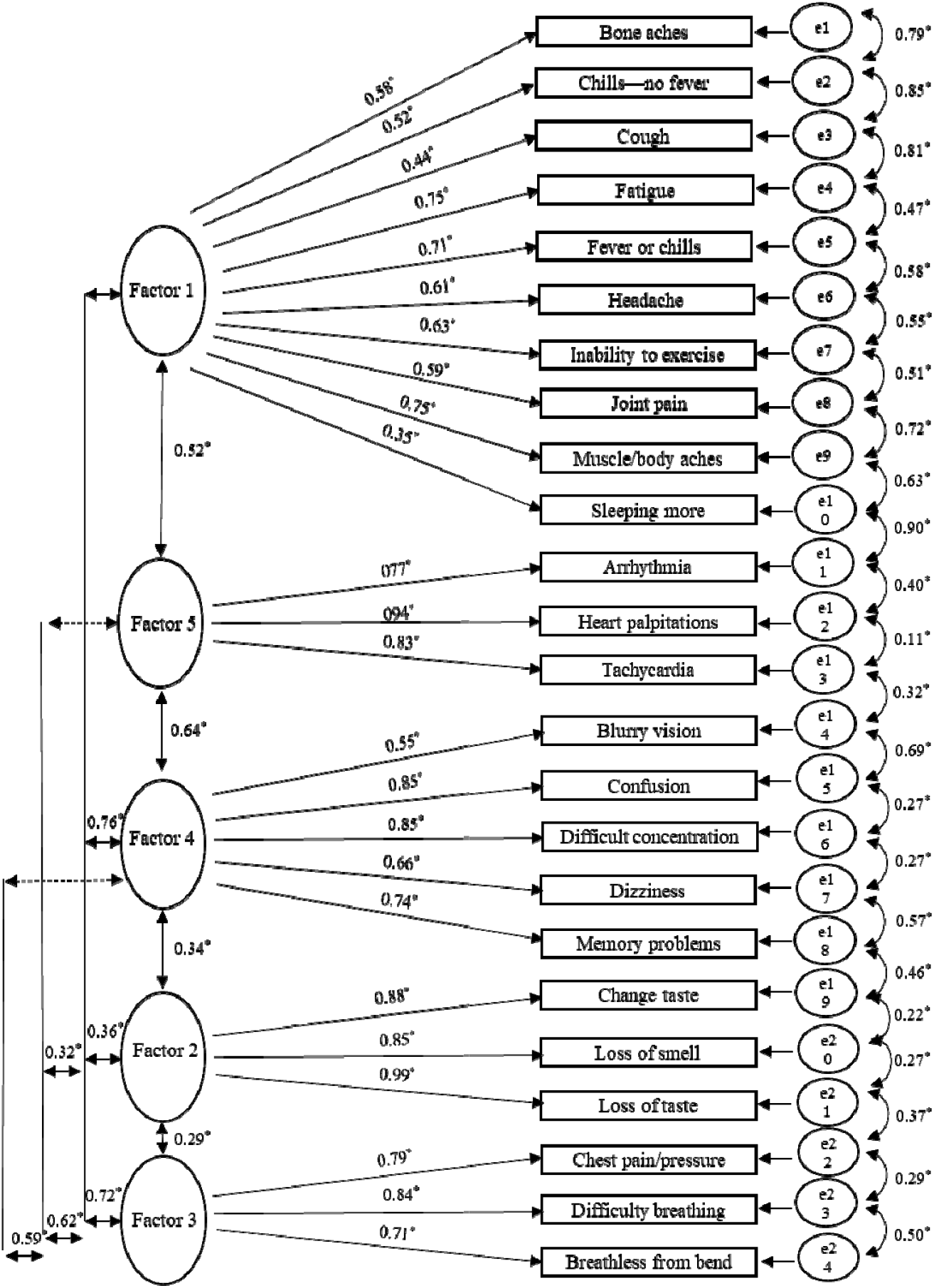
SEM Diagram PASC Confirmatory Factor Analysis. Initial SEM model demonstrating 5 factors with symptom loadings denoted. s1, Arrhythmia; s2, Blurry vision; s3, Bone aches in extremities; s4, Changed sense of taste; s5, Chills but no fever; s6, Confusion; s7, Cough; s8, Difficulty concentrating or focusing; s9, Dizziness; s10, Fatigue; s11, Fever or chills; s12, Headache; s13, Heart palpitations; s14, Inability to exercise or be active; s15, Joint pain; s16, Memory problems; s17, Muscle or body aches; s18, Partial or complete loss of sense of smell; s19, Partial or complete loss of sense of taste; s20, Persistent chest pain or pressure; s21, Shortness of breath or difficulty breathing; s22, Shortness of breath or exhaustion from bending over; s23, Sleeping more than usual; s24, Tachycardia.

### Confirmatory Factor Analysis Confirms Pattern of Symptom Onset

A five-factor structure was examined for validity with the second independent sample of 2,589 PASC survivors. Table 4 provides the goodness-of-fit for the 5-factor structure from the EFA. The initial model is the 5-factor model with all items retained from the EFA. A stepwise approach was then taken whereby paths from the stated items error terms were added sequentially. The final structure model was the best fitting model (*x*^2^ = 513.721, df= 207, p<0.01, TLI= 0.952, CFI= 0.964, RMSEA= 0.024) and the final model is shown in Figure 3.

**Figure 3:**
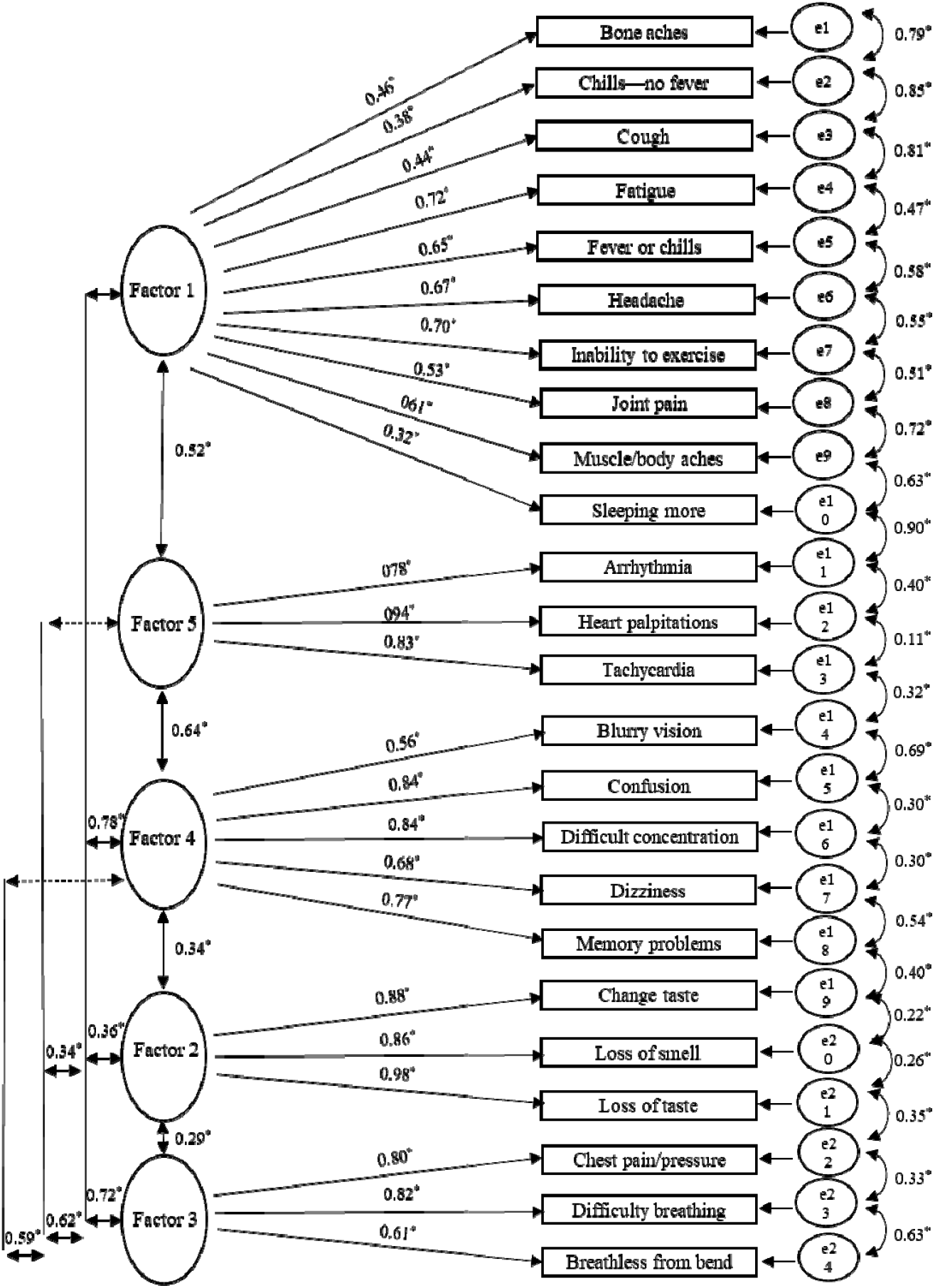
SEM Diagram of Non-PASC Confirmatory Factor Analysis. Final SEM model demonstrating 5 factors with symptom loadings denoted. s1, Arrhythmia; s2, Blurry vision; s3, Bone aches in extremities; s4, Changed sense of taste; s5, Chills but no fever; s6, Confusion; s7, Cough; s8, Difficulty concentrating or focusing; s9, Dizziness; s10, Fatigue; s11, Fever or chills; s12, Headache; s13, Heart palpitations; s14, Inability to exercise or be active; s15, Joint pain; s16, Memory problems; s17, Muscle or body aches; s18, Partial or complete loss of sense of smell; s19, Partial or complete loss of sense of taste; s20, Persistent chest pain or pressure; s21, Shortness of breath or difficulty breathing; s22, Shortness of breath or exhaustion from bending over; s23, Sleeping more than usual; s24, Tachycardia.

## Discussion

To our knowledge this study is one of the first nationwide studies conducted among PASC survivors to document a distinct and significant pattern of symptoms (symptom type and onset) among non-hospitalized persons with confirmed SARS-CoV-2 infection. Findings from this study warrant additional discussion and context to better understand their nursing implications.

We and others have described and characterized a broad range of symptoms associated with SARS-CoV-2 infection, as well as symptoms that persist for months and are a hallmark feature of PASC (3, 4, 9, 10, 13, 20). A limitation shared among many published studies is the use of checklists derived from clinician experience in managing COVID-19. Our initial understanding of symptoms associated with COVID-19 arose from our understanding that SARS-CoV-2 predominantly affected the respiratory system which limited initial symptom data. The depth and breadth of symptoms experienced among PASC survivors is just emerging. Indeed, an international study showed PASC survivors experience over 55 symptoms (13). Moreover, in our own work (3, 9), findings at illness presentation associated with the development of PASC is more extensive than the initial hallmark symptoms of SARS-CoV-2 infection, especially as asymptomatic persons are also at risk.

This study provides new information that points to a distinct and statistically significant pattern of PASC symptoms based on symptom type and symptom onset originally derived from unstructured patient reports. Additionally, this study adds to the science by providing a more granular assessment of PASC symptoms, early post-infection, than is has been measured other longitudinal work that commonly report within set timeframes (e.g., 4-week intervals). Our findings suggests that the evolution of PASC symptoms may follow a predictable pattern inclusive of a long duration (M days post-infection=105 days). This observation is troubling as it suggests that for some the sequalae of SARS-CoV-2 infection are ongoing and may be life-long. Further, this protracted length of illness suggests a major public health crisis is emerging and is affecting previously healthy and those with pre-morbid conditions alike. Finally, developing and implementing preventative interventions hinges upon understanding the course of the disease in the absence of any intervention. Given that PASC has no treatments or cure, this study documents the temporal order of symptoms during the evolution of the disease, in the absence of intervention, and provides the foundation for preventative and self-management strategies for PASC symptoms.

The confirmed factors (cold & flu like symptoms, change in smell and taste, dyspnea & chest pain, cognitive & visual symptoms, and cardiac symptoms) follow an arguably sequential pattern that could accompany progressive or persistent inflammation accompanied by endothelial activation. Other studies report elevated concentrations of a variety of cytokines (i.e., IL-6, and others) as well as persistent activation of immune cells for many weeks after SARS-CoV-2 infection (21-24). However, few studies have used biomarkers to correlate biological processes with symptom trajectories. This is an area ripe for exploration among nurse researchers.

A strength of this study is that it included mostly women aged 30-59 who identified as White. This is advantageous solely for conducting EFA and CFA because limiting sample collection from a wide range of populations is recommended to minimize the chance that factors present in one population will be obscured when pooled together with other populations. While this may limit the generalizability of the findings, it is nonetheless important because prior research has demonstrated that PASC affects women to a greater extent than men (3, 25) Further, this research serves as a basis for future comparisons between sex, races, ethnicities, and age groups to guide further research. This study asked PASC survivors to recall symptom type and their onset which is subject to recall bias unless symptoms were recorded via diary or tracking in real time by PASC survivors. However, viewing this weakness should be balanced with weaknesses of other methods of data collection such as clinician recorded symptoms (e.g. in the electronic health record) which has been shown to be discordant with patient reports (26, 27).

Ideally future work should be prospective and leverage multiple data sources in real time including patient report and triangulating data sources (e.g., wearable devices, patient report, clinical diagnostics, etc.…) that have been used in other studies. Additionally, prospective cohort data studies are particularly difficult to establish an emerging pandemic as pertinent constructs may not be known.

Second, the survey was conducted among individuals with symptoms following SARS-CoV-2 infection, thus excluding those who were asymptomatic. Moreover, most participants did not require hospitalization and/or supplemental oxygen delivery and would be considered to have mild-COVID-19. Most research in this area has been among hospitalized patients with COVID-19, limiting generalizability to those with less severe or asymptomatic initial presentation. (7, 10, 28). However, these data should not suggest that persons with asymptomatic infection do not develop PASC. Multiple studies suggest that those with asymptomatic SARS-CoV-2 infection are also at risk for long-term sequelae from COVID-19, although the incidence is unclear. In our work and others, data suggest that persons with PASC likely experience an evolution of symptoms that is unique from persons who do not develop PASC despite SARS-CoV-2 infection (3, 9, 28) Model validation utilizing a large sample of patients with SARS-CoV-2 who report prompt resolution of symptoms should be used to validate and understand if, and, or when a bifurcation in symptoms happens.

Third, this survey was conducted earlier in the COVID-19 pandemic when the alpha variant was dominant. Because the delta variant is far more contagious and symptom presentation differs slightly (29), it is unclear if the pattern reported here would be consistent between the two variants. This is an ongoing and active area of research for our team. Also, break through infections are occurring among vaccinated persons, and data suggest that vaccinated individuals are less likely to develop PASC following SARS-CoV-2 infection. However, additional studies are needed to assess PASC among the vaccinated, and should a new variant arise that minimizes the efficacy of current vaccines, the effect of breakthrough infections in the development of PASC would need to be re-evaluated.

Finally, there are potential implications for research and clinical practice (30). Nursing has an established history in symptom science and our collective expertise is needed at this time to address an unprecedented global health crisis. In addition to advancing the biological and molecular underpinnings of symptoms, Nursing is poised to rapidly re-tool and repurpose symptom management interventions to address symptoms among individuals with PASC. As the science develops around PASC, it may be possible to implement pharmacological and non-pharmacological interventions early during SARS-CoV-2 infection to mitigate potential long-term sequelae. An effective response to this global health crisis will require clinical and research collaboration among nurses to address the symptom burden afflicting the millions and rising numbers of individuals with PASC.

## Conclusion

PASC is a global health crisis that has emerged in response to the novel coronavirus, SARS-CoV-2. This study describes a distinct and statistically significant symptom pattern associated with PASC. We hope that this study will serve as a foundation upon which future studies can further characterize and understand PASC. As the pandemic continues, similar studies are needed to validate our findings and advance the understanding of PASC.

## Data Availability

All data produced in the present study are available upon reasonable request to the authors

## Acknowledgements

Thank you to Survivor Corps for mobilizing many thousands of COVID-19 survivors to participate in research to find a cure for COVID-19, and for being the epicenter of hope for so many. We would like to express our deep gratitude to the thousands of long-haulers who spent many hours taking this survey while suffering from the impacts of PASC. The answers we need to cure PASC will come from listening and learning from those who suffer from the disease.

We would also like to thank the Precision Health Initiative at Indiana University for their support on this project.

## Author contribution

All authors contributed to the conceptualization of the study, reviewed drafts, and provided final approval. CAD and MDP developed the analytic strategy, guided analysis, interpreted findings, drafted, and revised the manuscript. YH completed the analysis and AMR, and ND oversaw analysis and drafting of figures.

## Additional Information

The authors declare no competing interests.

## References

1. del Rio C, Collins LF, Malani P. Long-term Health Consequences of COVID-19. JAMA. 2020;324(17):1723–4.

2. Phillips S, Williams MA. Confronting Our Next National Health Disaster — Long-Haul Covid. N Engl J Med. 2021;385(7):577–9.

3. Yong SJ. Long COVID or post-COVID-19 syndrome: putative pathophysiology, risk factors, and treatments. Infect Dis (Lond). 2021 Oct;53(10):737–54.

4. Carfì A, Bernabei R, Landi F, Gemelli Against COVID-19 Post-Acute Care Study Group. Persistent Symptoms in Patients After Acute COVID-19. JAMA. 2020 Aug 11;324(6):603–5.

5. Carvalho-Schneider C, Laurent E, Lemaignen A, Beaufils E, Bourbao-Tournois C, Laribi S, et al. Follow-up of adults with noncritical COVID-19 two months after symptom onset. Clin Microbiol Infect. 2020 Oct 5.

6. Davis HE, Assaf GS, McCorkell L, Wei H, Low RJ, Re’em Y, et al. Characterizing long COVID in an international cohort: 7 months of symptoms and their impact. EClinicalMedicine. 2021 Aug;38:101019.

7. Halpin SJ, McIvor C, Whyatt G, Adams A, Harvey O, McLean L, et al. Postdischarge symptoms and rehabilitation needs in survivors of COVID-19 infection: A cross-sectional evaluation. J Med Virol. 2021 Feb;93(2):1013–22.

8. Tenforde MW, Kim SS, Lindsell CJ, Billig Rose E, Shapiro NI, Files DC, et al. Symptom Duration and Risk Factors for Delayed Return to Usual Health Among Outpatients with COVID-19 in a Multistate Health Care Systems Network - United States, March-June 2020. MMWR Morb Mortal Wkly Rep. 2020 Jul 31;69(30):993–8.

9. Huang Y, Pinto MD, Borelli JL, Mehrabadi MA, Abrihim H, Dutt N, et al. COVID Symptoms, Symptom Clusters, and Predictors for Becoming a Long-Hauler: Looking for Clarity in the Haze of the Pandemic. medRxiv. 2021 Mar 5.

10. Chopra V, Flanders SA, O’Malley M, Malani AN, Prescott HC. Sixty-Day Outcomes Among Patients Hospitalized With COVID-19. Ann Intern Med. 2020 Nov 11.

11. Lambert, N. J & Survivor Corps. COVID-19 “long-hauler” symptoms survey report. 2020.

12. Mandal S, Barnett J, Brill SE, Brown JS, Denneny EK, Hare SS, et al. ‘Long-COVID’: a cross-sectional study of persisting symptoms, biomarker and imaging abnormalities following hospitalisation for COVID-19. Thorax. 2020 Nov 10.

13. Davis HE, Assaf GS, McCorkell L, Wei H, Low RJ, Re’em Y, et al. Characterizing Long COVID in an International Cohort: 7 Months of Symptoms and Their Impact. medRxiv. 2020:2020.12.24.20248802.

14. Lambert, N., Survivor Corps, El-Azab, A.A., Ramrakhiani, N.S., Barisano, A., Yu, L., Taylor, K., Esperanca, A., Downs, C.A., Borelli, J.L., Chakraborty, R. & Pinto, M.D. The other COVID-19 survivors: timing, duration, and health impact of post-acute sequelae of SARS-CoV-2 (PASC) infection.. in review in review.

15. Costello, A & Osborne, J. Best practices in exploratory factor analysis: four recommendations for getting the most from your analysis. Practical Assessment Research and Evaluation. 2005;10(7):1–7.

16. Hatcher L. A step-by-step approach to using SAS for factor analysis and structural equation modeling. Cary, NC: SAS; 1994.

17. Cortina J. What is coefficeint alpha? An examination of theory and applications. Journal of Pediatric Psychology. 1993;32:288–96.

18. Bentler PM, Bonett DG. Significance tests and goodness of fit in the analysis of covariance structures. Psychol Bull. 1980;88(3):588–606.

19. Kline R. Principles and practice of structural equation modeling. The Guilford Press; 2004.

20. Bergquist SH, Partin C, Roberts DL, O’Keefe JB, Tong EJ, Zreloff J, et al. Non-hospitalized Adults with COVID-19 Differ Noticeably from Hospitalized Adults in Their Demographic, Clinical, and Social Characteristics. SN Compr Clin Med. 2020 Aug 14:1–9.

21. Doykov I, Hällqvist J, Gilmour KC, Grandjean L, Mills K, Heywood WE. ‘The long tail of Covid-19’ - The detection of a prolonged inflammatory response after a SARS-CoV-2 infection in asymptomatic and mildly affected patients. F1000Res. 2020 Nov 19;9:1349.

22. Lee CCE, Ali K, Connell D, Mordi IR, George J, Lang EM, et al. COVID-19-Associated Cardiovascular Complications. Diseases. 2021 Jun 29;9(3):47. doi: 10.3390/diseases9030047.

23. Proal AD, VanElzakker MB. Long COVID or Post-acute Sequelae of COVID-19 (PASC): An Overview of Biological Factors That May Contribute to Persistent Symptoms. Front Microbiol. 2021 Jun 23;12:698169.

24. Silva Andrade B, Siqueira S, de Assis Soares, W. R., de Souza Rangel F, Santos NO, Dos Santos Freitas A, et al. Long-COVID and Post-COVID Health Complications: An Up-to-Date Review on Clinical Conditions and Their Possible Molecular Mechanisms. Viruses. 2021 Apr 18;13(4):700. doi: 10.3390/v13040700.

25. Moreno-Pérez O, Merino E, Leon-Ramirez JM, Andres M, Ramos JM, Arenas-Jiménez J, et al. Post-acute COVID-19 syndrome. Incidence and risk factors: A Mediterranean cohort study. J Infect. 2021 Mar;82(3):378–83.

26. Donovan JL, Blake DR. Patient non-compliance: deviance or reasoned decision-making? Soc Sci Med. 1992 Mar;34(5):507–13.

27. Donovan JL. Patient decision making. The missing ingredient in compliance research. Int J Technol Assess Health Care. 1995;11(3):443–55.

28. Goërtz Ymj, Van Herck M, Delbressine JM, Vaes AW, Meys R, Machado FVC, et al. Persistent symptoms 3 months after a SARS-CoV-2 infection: the post-COVID-19 syndrome? ERJ Open Res. 2020 Oct 26;6(4):00542,2020. eCollection 2020 Oct.

29. Cella E, Benedetti F, Fabris S, Borsetti A, Pezzuto A, Ciotti M, et al. SARS-CoV-2 Lineages and Sub-Lineages Circulating Worldwide: A Dynamic Overview. Chemotherapy. 2021;66(1-2):3–7.

30. Pinto MD, Downs CA, Lambert N, Burton CW. How an effective response to post-acute sequelae of SARS-CoV-2 infection (PASC) relies on nursing research. Res Nurs Health. 2021 Aug 8.

